# “Odor identification task” actually measures global cognition – but with 70% error

**DOI:** 10.1101/2022.08.10.22277965

**Authors:** Rochelle E. Tractenberg, Futoshi Yumoto

## Abstract

**Objective:** To study the reliability and validity of an odor identification test.

**Methods:** The data come from an epidemiological cohort including 1146 non-Hispanic Caucasian, 86 Hispanic, and 12 other participants at the baseline visit (73.4% female). We tested the fit of each of three neurobiologically plausible models (validity) for responding on a 12-item odor identification task using confirmatory factor analyses (CFA); five model fit indices were assessed for each run. CFA testing fit *over time* (reliability) was planned for the measurement model that was found to fit across groups at the baseline visit. If a model was *<u>not</u>* found for the baseline visit, the test would be deemed “not invariant” over group, and not tested over time. In this case, we planned a *post hoc* Rasch analysis to further study test validity; and a multi-trait, multi-method analysis (MTMM) of the entire test battery to study reliability in terms of other, valid, cognitive and neuropsychological functional assessments.

**Results:** Nearly 70% of the variability in odor identification scores is *error*, a result that was replicated over four independent samples at the baseline visit. A core of 30% of “signal” from the task was identified over time (via Rasch modeling) but was explained fully by global cognition (replicated over time).

**Conclusions:** “Odor identification” as a construct cannot be reliably or validly measured over time or group. Multiple hypothesis-driven methods and replications show that this test provides no information that a global cognition score does not also (more validly and reliably) provide.

## Background

Recent analyses have suggested that impaired smell identification could identify both Alzheimer’s disease (AD) severity and the potential for the AD to progress rapidly (e.g., Velayudhan et al., 2013). Odor identification tasks have been suggested to represent olfactory cortical integrity or, increasingly, *general* cortical integrity (e.g., Wilson et al. 2009; see also Masurkar & Devanand, 2014). Hummel et al. (2011) report that that olfactory receptor neuron count decreases with age, but also that the threshold for smell detection can be unaffected by this change, supported by the findings of Li, Howard & Gottfried (2010), who report fMRI evidence that the observed deficit in odor identification in patients with Alzheimer’s disease is due to projections beyond olfactory cortex. Li et al (2010) reported that cognitive impairment (defined using Mini-Mental State (Folstein et al. 1975) scores) was not a factor in their observations of poorer odor identification performance in persons with AD vs. normal controls, while Velayudhan et al. (2013) reported that cognitive scores are significantly associated with both worse olfactory identification *and* greater decline in cognition.

Similar to Velayudhan et al. (2013) but in contrast to Li et al. (2010), Wilson et al. (2009) report that worse performance on an odor identification task was associated with greater AD neuropathology burden and not with episodic memory; this suggests that the observed relationship was *not* due to failures to recall odor names. The relationship between odor identification performance (tested just one time) and neuropathology burden post mortem is reported by Wilson et al. (2009) to be statistically significant, but no information has yet been published on how much of the variability in odor identification performance is *shared* with change in episodic memory (which has *also* been shown to be associated with neuropathology burden, see Wilson et al. 2007; Velayudhan 2015). In the 2009 Wilson, et al. study, the relationship between *change* in episodic memory and one-time odor identification performance is statistically significantly different from zero, and the strength of the relationship between odor identification test performance and later neuropathology burden was not affected when episodic memory was included in the model as a covariate.

High test-retest reliability (e.g., Doty et al. 1995; Doty 2007), combined with ease of administration and scoring, make a test with the potential to identify individuals with neurodegeneration early in the pathological process (i.e., before the emergence of frank cognitive symptoms) of great clinical and social interest. If validated as a proxy for neuropathology burden, AD severity, or the likelihood of a neuropathological process, an odor identification task would greatly improve the ability of primary care physicians and nurse practitioners outside of specialty clinics and AD research centers to detect and possibly also monitor important cognitive changes. However, although associations that are reported are statistically significant, analyses have not corrected for multiple comparisons - nor have any been replicated in independent samples; every study uses different methods and none of these is psychometrically appropriate. Further, the effect sizes have been small (as estimable from published papers). While test-retest reliability of odor identification tests have been reported for healthy controls, neither reliability nor validity, nor other psychometric characteristics have been published, although these features are critically important for documenting the utility of odor identification tests in the detection of AD or making clinical decisions about neuropathology using this information.

Moreover, odor identification involves naming, memory, and other cognitive functions that are affected directly by the Alzheimer’s disease pathology that the odor identification is hoped to help identify (see Li et al., 2010; Saive et al., 2014). The literature suggests that the roles of episodic memory and other cognitive functions in odor identification test performance have not yet been completely or accurately characterized; in their systematic review, Sun et al. (2012) concluded that more research was necessary to clarify and better estimate the relationships between odor identification tasks and AD. We therefore sought to study the psychometric and measurement properties of the odor identification task, to document its psychometric properties for use and utility in detecting Alzheimer’s disease or other neuropathology in primary care settings, and its validity for use over time.

Our research questions were: A) is there a single plausible measurement (latent variable) model of the odor identification test that will clarify and/or permit estimation of, the contributions of “smell” and “episodic memory”? And, B) if such a model exists, is it invariant over time and group (and race, as has been shown for a complex set of 17 neuropsychological tests in the same subjects by Barnes et al. 2016)?

## Measurement models

Our study focused on whether the odor identification task tests the olfactory system, or memory and naming, or some combination of these. To address the questions, we used latent variable modeling (LVM), described below. Specifically, applying an LVM approach articulates that if the odor identification test can be used as a meaningful proxy for neuropathology burden, then the odor identification items must meaningfully represent olfactory cortical, or general neuroarchitectural, integrity. This is evaluated statistically; causal latent variable modeling (see Bollen 1989; Grace, 2006) simultaneously estimates strengths and directions of associations and relationships among a set of variables that have been observed by modeling the moment structure of the data set. In this (causal) approach, the unobserved variable(s) for which we use the observed items as proxies are assumed to be <u>causing</u> the variation we observe in the performance on these items. Observed variables that share a common cause are called “indicators” of that common cause (see Mulaik, 2010, for discussion of emergent models).

When the unobserved (latent) common cause is modeled together with the observable indicators of this cause, the representation is called a “measurement model” because the indicators are the mechanisms by which the latent variable is measured (see Bollen 1989; Grace, 2006). As articulated by Bollen (1989), our challenge was to identify an appropriate measurement model for the odor identification items. The task itself is highly face valid; in order to answer any item correctly, a person clearly must be able to detect, and identify the source of, the odors that are represented on the task. However, if more than one skill can be used to correctly identify an odor (e.g., actual olfactory ability; memory; test-taking skill, etc.), then the measurement model that is most consistent with the observed performance would *also* need to represent these other skills – and their respective contributions can then be estimated. If a plausible measurement model cannot be identified, then the items can still be validated, but their measurement invariance (over groups or time) would be challenging to test and particularly, to interpret. That is, if we were able to document that the odor identification task requires a single (or main) underlying skill for performance on this test, it would support its use as a proxy for more robust (and time-, expertise-, and effort-intensive) cognitive/neuropathology assessments. On the other hand, if we found that memory or other cognitive skills, which are already included in cognitive batteries used to detect cognitive decline and neuropathology burden, represent a significant part of the model, then it would suggest that the odor identification task is not contributing uniquely to a clinician’s understanding of the patient’s neuropathology or cognitive function.

### Measurement Invariance

Given that an appropriate measurement model can be identified for the odor identification task, then *measurement invariance*, defined as statistical and theoretical evidence that measurement is consistent over groups and over time (or both) can be tested. Measurement invariance – while extremely important for characterizing tests that can be reliably utilized over time and across groups (e.g., male/female; old/young; Caucasian/African American, etc.) - is not a simple yes/no characterization (see, e.g., Horn & McArdle, 1992; Bollen & Curran, 2006; McArdle, 2009; Mulaik, 2010). In fact, invariance can be conceptualized as a continuum or hierarchy – and should be considered for time, if the test or instrument is to be utilized for the identification of change over time; or for groups, if performance on the test or instrument is to be compared across groups that might differ in their responses, level of performance, abilities, etc.; or *both* if groups are to be compared, or studied over time, using the same instrument.

Bollen & Curran (2006, pp. 254-57) specify that the most fundamental measurement invariance pertains to three features of the measurement model: model form, factor loadings, and intercepts. If the test is invariant in <u>model form</u>, the same indicators (observed variables or test items) will represent the same latent construct(s). If the test is invariant in its <u>factor loadings</u>, then given the same model form and scaling, the observed variables will relate to (load on) the latent variable(s) in the same way. If the test is invariant in <u>intercepts of the observed items</u> (indicator variables), then **the items (indicators) are functioning (representing the latent variable) in the same way**. The “sameness” referred to here can be with respect to time or group, or (preferably) both.

Measurement models that are shown to be invariant (i.e., can be replicated) across independent samples and over time improve science (Mulaik, 2010, pp. 283-4) as well as justifying inferences made over time and/or groups; however the documentation of invariance over time or groups also strengthens the validity and reliability of the instrument for any single administration (and its interpretation). Therefore measurement invariance is an important step in the creation and validation of any measurement instrument. For example, if a single-visit analysis suggest that the odor identification items measure both olfaction *and* memory/naming, then it is possible to control for memory/naming, and then test whether just the odor identification measurement features of the tests are invariant (over time, racial group, and both). Additionally, given a measurement model that fits the odor identification items well (Hu & Bentler, 1999), does every item on the test measure odor identification (and memory and naming, if these are also part of what the test measures) in the same way? Questions of measurement invariance cannot be answered without a measurement model that fits the data well.

## METHOD

### Subjects

The Memory and Aging Project (MAP) is an NIH-funded longitudinal cohort study that started in 1997, whose 1248 participants as of October 2012 contributed baseline data to these analyses. Both the study and the participants have been described extensively elsewhere (Bennett et al. 2012; Bennett et al. 2018). The Institutional Review Board of Rush University Medical Center approved this study (data available from https://www.radc.rush.edu/docs/studyPublications.htm?studyName=MAP). A brief descriptive summary of the sample is that there were 1146 non-Hispanic Caucasian, 86 Hispanic, and 12 other participants at the baseline visit (73.4% female). Table 1 shows the sample statistics. The MAP study is an ongoing epidemiologic cohort study, and at the time of these analyses, sufficient sample sizes to support the complex modeling we describe below were only available from the first three visits (baseline, visit 0, and one and two years after baseline, visits 1 and 2, respectively).

**Table 1.** Descriptive statistics at baseline.

|  | N | Mean | SD |
| --- | --- | --- | --- |
| Education (years) | 1260 | 14.38 | 3.29 |
| Age at baseline | 1260 | 79.43 | 7.91 |
| olfactory identification score | 1260 | 109.37 | 10.89 |
| global cognition score * | 1260 | 0.03 | 0.61 |
| Episodic Memory (EP) | 1260 | 0.052 | 0.74 |
| Semantic Memory (SE) | 1250 | 0.02 | 0.71 |
| Working Memory (WO) | 1260 | 0.01 | 0.78 |
| Perceptual Speed (PS) | 1249 | 0.01 | 0.83 |
| Visuospatial Abilities (PO) | 1239 | 0.01 | 0.81 |
| Notes: All values from baseline visit; scores are cognitive variables, derived from a battery described in Wilson et al. (2007); Barnes et al. (2014) and Bennett et al. (2012). These scores are derived (see Barnes et al. 2014) from Digits forward; Digits backward; Digits Ordering; Boston Naming Test; Category Fluency; Line orientation; Progressive Matrices; Story A Immediate Recall; Story A Delayed Recall; East Boston Memory Test; East Boston Delayed Recall; Word List Learning; Word List Recall; Word List Recognition; Symbol Digits Modality Test; Number Comparisons Test; Stroop, color naming test; Stroop, word reading test. |  |  |  |
| * These scores are normed so that 0 is the average score (see Barnes et al. 2014). |  |  |  |

### Instruments

The odor identification task in this data set is the12-item Cross-Cultural Smell Identification Test (CC-SIT) (Doty et al. 1996). The other instruments (Mini-Mental State Examination; Logical Memory Ia; Logical Memory IIa; Immediate story recall; Delayed story recall; Word List Memory (3 trials); Word List Recall; Word List Recognition; Complex Ideational Material; Boston Naming Test; Category Fluency (fruits, animals); National Adult Reading test; Digit Span Forward; Digit Span Backward; Digit Ordering; Symbol Digit Modalities Test; Number Comparison; Stroop word reading; Stroop color naming; Judgment of Line Orientation; Standard Progressive Matrices) are all discussed, with relevant references, by Bennett et al. (2012). These instruments were combined into cognitive factor scores reflecting Semantic Memory (SE); Visuospatial Abilities (PO); Perceptual Speed (PS); Episodic Memory (EP); Working Memory (WO) and global cognition (COG) (described in Barnes et al., 2016).

### Modeling

To address the question of measurement invariance (MI) in the smell test across groups and/or time, a measurement model was needed that could then be testable for its invariance. That is, the invariance question cannot be addressed until the *measurement* question is addressed. The first stage of analyses therefore addressed research question A): Is there a single plausible measurement (latent variable) model of the odor identification test that will clarify and/or permit estimation of, the contributions of “smell” and “episodic memory”? To answer this question, we compared the fits of three alternative models, shown in Figure 1, to the data.

**Figure 1.**
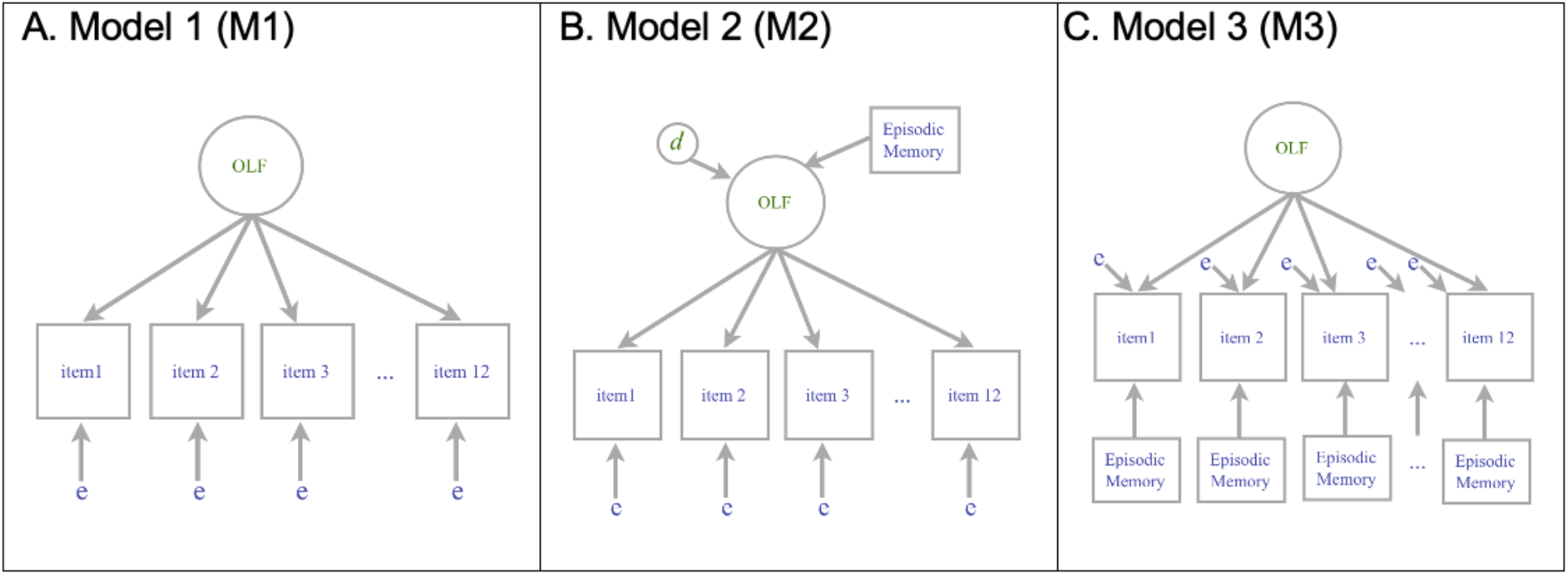
Three alternative measurement models of the 12-item odor identification test.

In Figure 1, circles represent latent (unobserved) variables and boxes/rectangles represent observed variables. The models are shown with only a few of the 12 observed (right/wrong) answers to the 12 items on the test, with latent “error” terms (e –shown without circles for simplicity) for each of the items. This simplifies the visual representation, but all models were actually based on all 12 items.

While theory and the literature suggest a one-factor causal model of odor identification task performance (M1), in two other models (M2, M3) we explicitly include contributions of episodic memory to odor identification, and memory is represented as an observed (not a latent) variable because of its derivation (see Barnes et al., 2016) and its inclusion in the dataset as a computed score for each participant. Each model was designed to represent plausible conclusions about the origins and face validity of the odor identification test and prior research on the relationships between odor identification and episodic memory. Each of these models was fit to the data at the baseline visit using a confirmatory method (described in the next section). We started with the baseline data because if a model does not fit a group at one visit, then it is not possible for that model to be measurement invariant over time for that group.

<u>Figure 1, Model 1 (M1)</u>: “null” model: This model is our “null” because it represents the simplest source of the face validity of the task and its general interpretation throughout the literature, namely, a test of odor identification arguably requires some, *primarily* olfactory, ability. If this model fits, it suggests that there is one (or one *primary*) latent construct, “olfactory ability” or possibly, “olfactory cortical integrity” (OLF), that is measured by the set of 12 smell items. If this model does not fit, then it suggests that there are other – significant - contributing skills or abilities that cause variability in smell item performance. Typically, the “null” model represents independence of the indicators (Bollen, 1989), but that (null) model has neither meaning nor interest in this context.

<u>Figure 1, Model 2 (M2)</u>. A multiple indicator **multiple causes** (MIMIC, Jöreskog and Goldberger, 1975) model, with one latent and one observed causal factor, where episodic memory contributes to the variability in the OLF factor and so, indirectly, to item performance. If this model fits the data well and better than the other alternatives, then the error term in this model with respect to the OLF factor (the disturbance term, “d”) will estimate how much variability in “odor identification ability” as measured by this test is NOT due to the factor labeled “Episodic Memory” -i.e., is specific to the factor labeled “olfaction”. The item-level error terms are shown as a single oval for items but are estimated individually per item. Model 2, if it fits best & well, implies that performance on the smell test is at least partly a function of top-down processing. This has important clinical implications for the interpretation of smell test performance in screening for AD/MCI, because it suggests that performance is not simply a representation of the status of olfactory cortex (as a proxy for overall brain health, see Saive et al. 2014).

<u>Figure 1, Model 3 (M3)</u>. Two independent (residualized) contributions to variability in the smell items performance; one latent factor for olfaction (OLF) and one observed episodic factor score contributing causally to each item. In this model, the error terms are all on the items (shown in a single oval but estimated for each item) but not on the OLF factor (i.e., no disturbance term). If this model fits the data, then these error terms estimate how much variability in response is due to *neither* smell nor episodic memory, i.e., the “measurement error” for these items in this model. The explicit modeling of the *independence* contributions of episodic memory and olfactory ability to performance on the test items represents smell as if it is a primary cortical area, which it is. The contribution of memory to the response is then modeled separately, which is physiologically plausible.

The neuroanatomical literature suggests that primary olfactory cortex does not receive much top-down processing, and olfactory cortical outputs do not pass through thalamus or other relay stations (Saive, et al., 2014). Therefore, it is plausible that olfactory ability (to the extent it is reflected by performance on this test) may be independent of at least some part of smell *naming*, and M3 reflects that explicitly. By contrast, M2 explicitly models the opposite. In Model 1 (M1), the independence is not modeled at all, and if M1 fits the data well and best, contributions of episodic memory would be undifferentiated in the “error” terms that capture all of the variability in each item that is not directly due to (caused by) variability in the olfactory factor.

## ANALYSIS

The analyses were planned to be completed in two stages. In the measurement modeling stage, we ran confirmatory factor analysis (CFA), for each of the three alternative models, in roughly one quarter of the over 1200 participants with this test data at their baseline visit. This would permit the later stage multi-group CFAs on (validation) samples from the same cohort at baseline, strengthening our support for the “winning” measurement model of the odor identification test. If just one, or if *neither* stage, was successful, we planned hypothesis-driven post hoc analyses of <u>reliability</u> (as model misfit through a residuals analysis and Rasch modeling) and <u>validity</u> (multi-trait, multi-method analysis). Interpretation of these analyses did not depend on specific inference tests, so no corrections for multiple comparisons were planned (or completed), and a power calculation was not necessary.

### Measurement modeling stage

Confirmatory factor analyses (CFA) were carried out using Mplus 7.0 (Muthen & Muthen, 2012). These analyses tested the correspondence of the measurement model alternatives (M1-M3) shown in Figure 1 above with 25% random splits of the data at the baseline visit. Five indices, describing the appropriateness of the model given the data (details below) along multiple dimensions of the model-data fit (i.e., not simply a chi-square statistic for model fit) were assessed for each run: Satorra-Bentler model chi square (*χ*^2^); general robust model fit statistic (general data-model fit), with the associated p-value for the degrees of freedom; robust Akaike’s Information Criterion (AIC; fit of the model to data in hypothetical replications; the lower, the better; CFI: Robust Comparative fit index (incremental model fit relative to independence); the closer to 1.0 the better, acceptable models have CFI>.96; SRMR: standardized root mean square residuals (mean absolute value of the covariance residuals), the smaller (and <0.09) the better; and RMSEA: Robust root mean square error of approximation (error in approximation of the data); the closer to zero (and positive) the better, acceptable models have an upper bound on the 90% CI <0.06 (Hu & Bentler, 1999).

Model fit statistics were compiled in order to determine which of the three plausible alternative measurement models had good (all fit indices in acceptable range), and the best (lowest AIC value), fit. The best fitting, “winning”, model would be the one where all fit indices are consistent in indicating the best (most desirable) value.

In addition to examining and comparing performance of fit indices outlined above, the residuals (comparing the observed and model-implied variance-covariance and other moment structure matrices), and modification indices were examined for each model run. The residuals indicate, where relevant, those indicators (items) for which a model does not fit well, while the modification indices provide a relative comparison of the misfit of the hypothesized relationships in a given model. Reasons for misfit might include covariances between items that are not in the (original) model. We planned to use the residuals and modification index values to modify the alternatives in Figure 1, in order to increase the likelihood of having a single measurement model for the smell test items to pursue in the invariance testing stage.

The use of independent replications of modeling results by randomly splitting the 1200+ person cohort was planned to strengthen conclusions; this approach maintains sufficient sample size (minimum of 250 observations per model with 12 indicators; see Wolf et al., 2013) while also providing critical replication of findings.

### Invariance testing stage

Multi-group confirmatory factor analyses (MG-CFA) using Mplus 7.0 (Muthen & Muthen, 2012) were planned for the “winning” measurement model from the first analysis stage. First, taking the remaining cohort (N=927) after the random split (N=315) for the measurement modeling stage and randomly dividing it in thirds, we were able to create three independent cross validation groups (n=315 each) at baseline. We planned a series of tests of MI to test for: invariance over three visits within group; invariance across these three groups by visit; and invariance over both features together.

In our study, we planned to fit a fully constrained version of the winning model (from stage one) first, relaxing constraints iteratively from fully constrained (factor structure, indicator loadings, and indicator intercepts each constrained equal), to measurement constrained (i.e., releasing constraints on the intercepts, but not loadings or factor structure), to structural (i.e., same items loading on same factors without the values of the loadings or intercepts constrained), to fully *unconstrained* (all factors, loadings, and parameters in the set of models estimated separately). Fit of the model to the data would be defined using the same fit statistics as in stage 1 (Hu & Bentler, 1999).

### Planned post-hoc analyses

If none of our alternative models (Figure 1) fit the odor identification task items in the measurement modeling stage, then the answer to research question (A) “is there a single plausible latent variable model of the odor identification test that will clarify and/or permit estimation of, the contributions of “smell” and “episodic memory”?” would be “none of those in Figure 1”. A failure of measurement model fitting would also make it impossible to address research question (B) “if such a model exists, is it invariant over time and group?” However, failures in either or both stage would still leave analysis options: hypothesis-driven post hoc analyses of reliability through Rasch modeling (model misfit and residuals analysis) and validity (multi-trait, multi-method analysis).

In the event that none of our plausible measurement models fit the data, and no plausible/theoretically reasonable modifications were suggested by the evaluation of the residuals and modification indices, we planned to explore two dimensions of the odor identification items. First, hypothesis-driven post hoc analysis of <u>reliability</u> would be achieved through estimation of the misfit of a Rasch model to the data, with a residuals analysis. In this approach we would be interested in whether a unidimensional (Rasch) model of the data, demonstrated to be inappropriate by failures of stage one analyses (i.e., M1 is unidimensional), would nevertheless yield “interpretable” results about the odor identification test and its reliability. The Rasch model assumes a unidimensional, causal measurement model (Rasch, 1960/1980). Further, the fit statistics for modern measurement models such as the Rasch, “infit” and “outfit” values, which represent squared standardized response residuals and thereby provide important information about the reliability of a single “summary” score –e.g., a total or sum of item scores - derived from the items thus modeled. Infit and outfit are commonly employed to characterize the fit (or misfit) of the data to the Rasch model (although see Kreiner & Christensen, 2011, who argue that the *distributions* of these indicators do not function in ways that support their systematic use and interpretation). Second, further information about psychometric <u>validity</u> of the odor identification test could be derived from a multi-trait, multi-method analysis involving the entire cognitive battery. In this hypothesis-driven analysis we would determine whether any patterns of associations could be suggestive of areas of validity, or future measurement modeling for this test. These insights can be ascertained from a multi-trait multi-method (MTMM, Campbell & Fisk, 1956) approach to estimating the relative strengths of associations among all of the outcomes that this rich cohort has available.

The Rasch modeling would be achieved using Winsteps 3.81.0 (Linacre, 2014) with particular focus on the psychometric evaluation of individual items through the examination of item parameters and fit statistics and residual factor structure through the principal component analysis of residuals. In the case that we carried out this modeling, we would have known that the single factor model did not fit the odor identification data given the results of the measurement modeling stage.

The construction of a multi-trait, multi-method (MTMM) analysis of the set of outcomes being collected in this longitudinal study cohort entails a simple (Pearson) correlation matrix arranged according to Campbell & Fisk (1959, p. 82). Because the odor identification task was the only assessment of this particular trait in a battery with multiple measures of both general and specific cognitive functioning, we were most interested in testing whether the correlations between the odor score and cognitive scores, which are arguably tests designed to get at different traits, were *lower* than correlations between the various independent tests of general and specific cognitive functioning. We would also seek to determine whether the same patterns were observed among these tests over time.

## RESULTS

Table 1 gives descriptive statistics for the cohort at baseline.

### Measurement modeling

Table 2 shows the fits of the three alternative models for the first randomly split subset.

**Table 2.** Fit indices for three models fit to one randomly split subgroup (n=315) at baseline.

|  | Model (see Figure 1) |  |  |
| --- | --- | --- | --- |
| Fit Index: how did the model fit the data? | Model 1 | Model 2 | Model 3 |
| $\chi^2$ (df, p) | 461.5 (df=282, p<0.001) | 533.4 (df=329, p<0.001) | 511.4 (df=318, p<0.001) |
| AIC | 35902 | 35367 | 35367 |
| CFI | 0.626 | 0.643 | 0.662 |
| SRMR | 0.063 | 0.065 | 0.063 |
| RMSEA | 0.045 | 0.045 | 0.044 |
| Notes: AIC = Akaike's Information Criterion; CFI=comparative fit index; SRMR= standardized root mean square residuals; RMSEA = root mean square error of approximation. |  |  |  |

All three alternative models – including the one factor-were poor fits to these data. None of the fit indices was in the acceptable range. These results demonstrate that, in spite of the intuitive sense of unidimensionality of the odor identification test, the test does not have a single (main) factor underlying it. Moreover, M2 and M3 in Figure 1 each represent plausible interpretations of published studies and data characterizing relationships between this test and memory performance; neither of those models fit the data either. We created a second 25% random split of the remaining sample and re-ran the models with the same results (poor fit of all models to each subsample; data not shown). Examination of the residuals for each model showed that none of the items exhibited particularly bad fit, the inclusion of the episodic memory scores (M2, M3) was also not a source of particular misfit, and none of the suggested modifications to any one of the models (for either group) had any rational interpretation. For example, if one model had a modification index that suggested that two items’ errors should be correlated, this was not observed in the other models or in the other subsample. All of the evidence suggested that these models – including the single factor “null” model – were bad fits to these data, including our independent replication, suggesting that the odor identification test does not have measurement invariance properties that are consistent with published literature about this test.

Because the measurement modeling stage failed, the multi-group CFA were not done (stage 2), and we moved on to our hypothesis-driven post hoc analyses.

### Post Hoc Analyses: Rasch modeling

This unidimensional model was fit to data at each of the first three visits for the entire sample (n=1260 (visit 0); n=200 (visit 1); n=181 (visit 2)). As noted, the fit of a Rasch model is established through both model- and item-level statistics. Our Rasch modeling results (Tables 3-4) show clear failures of the items to fit this unidimensional model structure.

**Table 3.** Model-level analysis of Rasch model fit: Principal components analysis of *residuals* of Rasch model of 12-item odor identification test over time (full sample) showing percent of variance in total scores explained by first three orthogonal dimensions extracted from residuals.

|  | % Variance explained by dimensions discovered in Rasch model residuals |  |  | Total variance in residuals explained by first 3 dimensions |
| --- | --- | --- | --- | --- |
| Time | Dimension 1 | Dimension 2 | Dimension 3 |  |
| Baseline | 32.8 | 7.7 | 6.6 | 47.1% |
| Year 1 | 29.7 | 9.2 | 8.5 | 47.4% |
| Year 2 | 30.0 | 8.9 | 8.6 | 47.5% |
| Notes: Residuals of a Rasch model should have no discernable structure. Finding dimensions in the residuals of a Rasch model is evidence against unidimensional structure *and* invariance of the model over time/groups. The first dimension recovered from PCA of residuals is the “main” dimension; the next two dimensions are not strong but are consistently found over time –as is the “main” dimension. The main dimension accounts for just 30% (or so) of the variability of the test score at any one visit. |  |  |  |  |

**Table 4.** Item level-analysis of Rasch model fit: Identification of items on the 12 item odor identification task that do not fit the unidimensional Rasch model in the full sample (N=1260) at each visit.

|  | Baseline |  | T1 |  | T2 |  | Visits (of 3)<br>that the item<br>does not fit |  |
| --- | --- | --- | --- | --- | --- | --- | --- | --- |
| Item | Infit | Outfit | Infit | Outfit | Infit | Outfit | Infit | Outfit |
| 1 |  |  | x |  |  |  | 1 |  |
| 2 |  | x |  | x |  |  |  | 2 |
| 3 |  | x | x | x | x | x | 2 | 3 |
| 4 |  | x |  | x |  | x |  | 3 |
| 5 |  |  |  | x |  | x |  | 2 |
| 6 |  |  |  | x |  |  |  |  |
| 7 |  | x |  | x |  |  |  | 2 |
| 8 |  |  |  |  |  |  |  |  |
| 9 |  |  |  |  |  |  |  |  |
| 10 |  | x |  | x |  | x |  | 3 |
| 11 |  |  |  | x |  | x |  | 2 |
| 12 |  |  |  | x |  |  |  | 1 |
| Total |  | 5 | 2 | 9 | 1 | 5 | 3 | 18 |

Note: The fit of a Rasch model to data can be explored as a function of items. If particular items cause failures to fit, they can be further examined or eliminated. If no particular item causes failures to fit, the data simply do not fit the Rasch model, and hypotheses of both unidimensionality and invariance are rejected.

Residuals of a Rasch model should have no discernable structure. Finding dimensions in the residuals of a Rasch model is evidence *against* unidimensional structure and invariance of the model over time/groups. A principal components analysis of the Rasch model residuals should show no structural features (no factors).

Table 3 shows two key results about the odor identification test: first, the Rasch model explained about 30% of the total test score variance at each visit; this suggests that, while not essentially unidimensional, there may be one main dimension of the test, but it only captures 30% of the variability in the total test score. That means that <u>70% of the total score is error</u> – this 30% does <u>not</u> represent a core factor. The second observation is that there are two additional dimensions, capturing 7-9% and 6-8% of remaining variance, respectively, that are also consistently observed over time.

Rasch model results can also be investigated at the item level, by exploring the fit of each item with the assumptions and requirements of the model. Rasch modeling assumes essential unidimensionality, and failures are identified by items with fit statistics that fall outside the plausible range of item infit and outfit values (.8 – 1.2; see Wright & Linacre, 1994). The item-level fit results for the Rasch model at each of three annual visits are presented in Table 4.

Table 4 shows that there are two reasons to reject unidimensionality of the odor identification task: at least 40% of the items do not fit the Rasch model at any given visit; but even more problematic for this test and its interpretability is that the five items that failed to fit at baseline and those failing at the 2^nd^ annual visit are *different*. That is, the failure to fit is not due to the functioning of a few specific items, consistently over time. Different items fail to fit the Rasch model at different times, suggesting the items do not function predictably or reliably. Therefore, hypotheses of both unidimensionality and invariance are rejected for this test.

### Post Hoc Analyses: MTMM analysis

Table 5 presents the MTMM matrix showing correlations among the scores with each other and over time.

**Table 5.** Multi-trait Multi-Method matrix for odor identification task with cognitive battery scores: full sample, by visit.

|  |  | Baseline |  |  |  |  |  |  | Time 1 |  |  |  |  |  |  | Time 2 |  |  |  |  |  |  |
| --- | --- | --- | --- | --- | --- | --- | --- | --- | --- | --- | --- | --- | --- | --- | --- | --- | --- | --- | --- | --- | --- | --- |
|  |  | Smell | EP | SE | WO | PS | PO | COG | Smell | EP | SE | WO | PS | PO | COG | Smell | EP | SE | WO | PS | PO | COG |
| Baseline | Smell | 1 | 0.4 | 0.4 | 0.3 | 0.4 | 0.2 | 0.4 | 0.8 | 0.3 | 0.4 | 0.2 | 0.4 | 0.2 | 0.4 | 0.7 | 0.3 | 0.3 | 0.2 | 0.3 | 0.1 | 0.3 |
|  | EP | 0.4 | 1 | 0.6 | 0.5 | 0.5 | 0.3 | 0.9 | 0.4 | 0.9 | 0.6 | 0.3 | 0.6 | 0.3 | 0.8 | 0.4 | 0.9 | 0.7 | 0.6 | 0.6 | 0.3 | 0.9 |
|  | SE | 0.4 | 0.6 | 1 | 0.6 | 0.7 | 0.5 | 0.8 | 0.4 | 0.6 | 0.9 | 0.5 | 0.7 | 0.6 | 0.8 | 0.4 | 0.6 | 0.8 | 0.5 | 0.6 | 0.4 | 0.8 |
|  | WO | 0.3 | 0.5 | 0.6 | 1 | 0.5 | 0.4 | 0.7 | 0.2 | 0.4 | 0.5 | 0.8 | 0.5 | 0.3 | 0.6 | 0.1 | 0.4 | 0.4 | 0.8 | 0.5 | 0.3 | 0.6 |
|  | PS | 0.4 | 0.5 | 0.7 | 0.5 | 1 | 0.5 | 0.8 | 0.3 | 0.5 | 0.6 | 0.5 | 0.9 | 0.5 | 0.8 | 0.3 | 0.5 | 0.6 | 0.5 | 0.8 | 0.4 | 0.7 |
|  | PO | 0.2 | 0.3 | 0.5 | 0.4 | 0.5 | 1 | 0.6 | 0.1 | 0.1 | 0.4 | 0.3 | 0.4 | 0.7 | 0.4 | 0.2 | 0.3 | 0.4 | 0.2 | 0.3 | 0.6 | 0.4 |
|  | COG | 0.4 | 0.9 | 0.8 | 0.7 | 0.8 | 0.6 | 1 | 0.4 | 0.8 | 0.8 | 0.6 | 0.8 | 0.5 | 0.9 | 0.4 | 0.8 | 0.8 | 0.7 | 0.7 | 0.5 | 0.9 |
| Time 1 | Smell | 0.8 | 0.4 | 0.4 | 0.2 | 0.3 | 0.1 | 0.4 | 1 | 0.5 | 0.5 | 0.3 | 0.4 | 0.3 | 0.5 | 0.7 | 0.1 | 0.2 | 0.1 | 0.2 | 0.1 | 0.2 |
|  | EP | 0.3 | 0.9 | 0.6 | 0.4 | 0.5 | 0.1 | 0.8 | 0.5 | 1 | 0.7 | 0.4 | 0.6 | 0.3 | 0.9 | 0.2 | 0.9 | 0.7 | 0.7 | 0.6 | 0.1 | 0.8 |
|  | SE | 0.4 | 0.6 | 0.9 | 0.5 | 0.6 | 0.4 | 0.8 | 0.5 | 0.7 | 1 | 0.6 | 0.7 | 0.5 | 0.9 | 0.4 | 0.7 | 0.9 | 0.5 | 0.5 | 0.6 | 0.8 |
|  | WO | 0.2 | 0.3 | 0.5 | 0.8 | 0.5 | 0.3 | 0.6 | 0.3 | 0.4 | 0.6 | 1 | 0.5 | 0.4 | 0.7 | 0.1 | 0.6 | 0.5 | 0.8 | 0.6 | 0.4 | 0.7 |
|  | PS | 0.4 | 0.6 | 0.7 | 0.5 | 0.9 | 0.4 | 0.8 | 0.4 | 0.6 | 0.7 | 0.5 | 1 | 0.4 | 0.8 | 0.5 | 0.6 | 0.8 | 0.6 | 0.9 | 0.6 | 0.8 |
|  | PO | 0.2 | 0.3 | 0.6 | 0.3 | 0.5 | 0.7 | 0.5 | 0.3 | 0.3 | 0.5 | 0.4 | 0.4 | 1 | 0.6 | 0.1 | 0.4 | 0.7 | 0.3 | 0.4 | 0.8 | 0.6 |
|  | COG | 0.4 | 0.8 | 0.8 | 0.6 | 0.8 | 0.4 | 0.9 | 0.5 | 0.9 | 0.9 | 0.7 | 0.8 | 0.6 | 1 | 0.3 | 0.9 | 0.9 | 0.8 | 0.8 | 0.5 | 1 |
| Time 2 | Smell | 0.7 | 0.4 | 0.4 | 0.1 | 0.3 | 0.2 | 0.4 | 0.7 | 0.2 | 0.4 | 0.1 | 0.5 | 0.1 | 0.3 | 1 | 0.5 | 0.5 | 0.4 | 0.5 | 0.4 | 0.6 |
|  | EP | 0.3 | 0.9 | 0.6 | 0.4 | 0.5 | 0.3 | 0.8 | 0.1 | 0.9 | 0.7 | 0.6 | 0.6 | 0.4 | 0.9 | 0.5 | 1 | 0.7 | 0.6 | 0.7 | 0.5 | 0.9 |
|  | SE | 0.3 | 0.7 | 0.8 | 0.4 | 0.6 | 0.4 | 0.8 | 0.2 | 0.7 | 0.9 | 0.5 | 0.8 | 0.7 | 0.9 | 0.5 | 0.7 | 1 | 0.6 | 0.7 | 0.7 | 0.9 |
|  | WO | 0.2 | 0.6 | 0.5 | 0.8 | 0.5 | 0.2 | 0.7 | 0.1 | 0.7 | 0.5 | 0.8 | 0.6 | 0.3 | 0.8 | 0.4 | 0.6 | 0.6 | 1 | 0.6 | 0.4 | 0.8 |
|  | PS | 0.3 | 0.6 | 0.6 | 0.5 | 0.8 | 0.3 | 0.7 | 0.2 | 0.6 | 0.5 | 0.6 | 0.9 | 0.4 | 0.8 | 0.5 | 0.7 | 0.7 | 0.6 | 1 | 0.5 | 0.8 |
|  | PO | 0.1 | 0.3 | 0.4 | 0.3 | 0.4 | 0.6 | 0.5 | 0.1 | 0.1 | 0.6 | 0.4 | 0.6 | 0.8 | 0.5 | 0.4 | 0.5 | 0.7 | 0.4 | 0.5 | 1 | 0.6 |
|  | COG | 0.3 | 0.9 | 0.8 | 0.6 | 0.7 | 0.4 | 0.9 | 0.2 | 0.8 | 0.8 | 0.7 | 0.8 | 0.6 | 1 | 0.6 | 0.9 | 0.9 | 0.8 | 0.8 | 0.6 | 1 |

NOTES: N=1239; BL = baseline visit; Y1 = first annual visit (one year after baseline); Y2 = second annual visit (two years after baseline). Smell = total score on olfactory identification test; Abbreviations: SE= Semantic Memory; PO = Visuospatial Abilities; PS=Perceptual Speed; EP= Episodic Memory; WO = Working Memory.

As noted, the entries in each cell in the MTMM matrix in Table 5 are the Pearson Correlation coefficients representing the association between the tests (row/column). Squaring any cell entry yields the shared variance between total scores on those two tests at baseline (visit 0) and at one and two years after baseline (visit 1, visit 2). Similar to the result in Table 3, Table 4 also shows that there is a “core” of variability (roughly 30%), which is identified by the (squared) correlation coefficient between the global cognitive and smell identification total scores in each of three annual visits. None of the other test scores is correlated as highly with the odor score.

The similarity in the Rasch and MTMM model results – identifying a roughly 30% “core” over time-suggested an unplanned *post hoc* analysis: The MTMM result suggests that the 30% core that was found across visits in the Rasch modeling might actually be due to global cognition. We tested this hypothesis by simple linear regressions, by visit, after partialling out the contribution of global cognition. We regressed each residualized variable on the residualized smell test total score, and then recomputed the correlations between the residualized versions of all scores in the test battery, The results are shown in Table 6.

**Table 6.** Reduced-dimension Multi-trait Multi Method table (correlations) for the residualized smell scores with global cognition partialled out (bold) of all cognitive scores

|  | Zsmell BL | Zsmell Y1 | Zsmell Y2 |
| --- | --- | --- | --- |
| zcog_ep BL | 0.05 | 0.01 | -0.03 |
| zcog_se BL | 0.00 | 0.01 | -0.02 |
| zcog_wo BL | -0.07 | -0.09 | -0.22 |
| zcog_ps BL | 0.01 | -0.10 | -0.02 |
| zcog_po BL | -0.08 | -0.07 | 0.00 |
| globcog BL | **0.00** | -0.05 | -0.07 |
| zcog_ep Y1 | 0.02 | 0.02 | -0.25 |
| zcog_se Y1 | 0.06 | 0.05 | 0.00 |
| zcog_wo Y1 | -0.08 | -0.12 | -0.27 |
| zcog_ps Y1 | 0.02 | -0.01 | 0.10 |
| zcog_po Y1 | 0.01 | -0.02 | -0.14 |
| globcog Y1 | 0.01 | **0.00** | -0.16 |
| zcog_ep Y2 | -0.05 | -0.33 | 0.01 |
| zcog_se Y2 | -0.10 | -0.24 | 0.06 |
| zcog_wo Y2 | -0.16 | -0.35 | -0.09 |
| zcog_ps Y2 | -0.05 | -0.17 | 0.01 |
| zcog_po Y2 | -0.10 | -0.13 | 0.00 |
| globcog Y2 | -0.09 | -0.32 | **0.00** |

Notes: N=1239; Smell = total score on olfactory identification test; Abbreviations: SE= Semantic Memory; PO = Visuospatial Abilities; PS=Perceptual Speed; EP= Episodic Memory; WO = Working Memory.
“z” in the variable name indicates that it is the residualized value (after regression with global cognition), i.e., these correlations are between each test score after the contribution of “global cognition” to each score was partialled out of each score.
BL = baseline visit; Y1 = first annual follow up; Y2 = second annual follow up.
Bold correlations indicate how global cognition (“globcog”) was partialled out of these scores. Cognitive scores here are derived from a battery described in Wilson et al. (2007); Barnes et al. (2014) and Bennett et al. (2012) that includes Digits forward; Digits backward; Digits Ordering; Boston Naming Test; Category Fluency; Line orientation; Progressive Matrices; Story A Immediate Recall; Story A Delayed Recall; East Boston Memory Test; East Boston Delayed Recall; Word List Learning; Word List Recall; Word List Recognition; Symbol Digits Modality Test; Number Comparisons Test; Stroop, color naming test; Stroop, word reading test.

In Table 6 it can be seen that when global cognition is partialled out of the other scores, there is *no* association between the residualized smell scores over time; the 30% “core” is eliminated. There are also no meaningful correlations between odor identification and the other residualized test scores across the MTMM matrix over time. This result supports the conclusion that the construct that was consistently identified in the Rasch modeling over time is actually global cognition, and not smell.

## DISCUSSION

These results, replicated across independent groups of sufficiently large sizes, show clear failures of the odor identification test to: a) fit a one-factor model; and b) fit two biologically-plausible alternative models. It might be tempting to attribute these failures to the models that we chose. This interpretation would be consistent with the high face validity of the test; and while the models that we studied were each consistent with some previous findings involving the constructs that are represented in the data we analyzed, the fact that none of *these* particular plausible alternatives fit the data is not especially important with respect to the utility of the (or any) odor identification test. More critical is the fact that a one-factor model did not fit any group, and that when we were able to identify a single factor, it a) explained just 30% of the total variability in test scores; and b) was fully eliminated when controlling for global cognitive functioning. Together, these results strongly suggest that the apparent face validity of the odor identification task is a misperception. Yeshurun & Sobel (2010) suggested that olfaction is “mistakenly” described as multi-dimensional; they argue that it is actually “nearly unidimensional” – and our results very concretely (and with independent replications) refute that claim.

When performance on an odor identification test has been found to be significantly associated with cognitive test performance, even if that association is “above and beyond” (after controlling for) cognitive test scores, that *would be expected* because between 2-25% of the variability in the smell test is *shared* with other cognitive tests and in large samples, even small correlations will reach statistical significance. Only about 30% of the variance in performance on the smell test itself at any time is actually due to the single core factor that we identified in every group at every timepoint; and between 2% and 25% of that “core” variability is actually conflated with, or possibly due to, variability in the functioning of these other (not-olfactory) factors. Furthermore, 75%-98% of the variability in odor identification is NOT shared with these other (non-olfactory) factors, so even if the remaining variability in performance is all noise, it is highly likely that associations that are significantly different from zero would be observed, for example, between odor perception and neuropathology – even after controlling for other variables.

In their systematic review, Sun et al. (2012) concluded that more research was necessary to clarify and better estimate the relationships between odor identification tasks and AD. Olofsson (2014) proposed a complex model of “olfactory behavioral decisions” that includes multiple paths from primary olfactory cortex detection to a decision on an identification task; Karunanayaka et al. (2014) also describe a highly complex system of networks (see also Li et al. 2010; Bowman et al. 2012; Wilson et al. 2014;Howard et al. 2016; Fournel et al. 2016) Any of the features of these models could be slowed, eliminated, or impaired to different degrees by aging and/or neuropathological processes, which could explain our failure to find a single latent variable model for performance on this task: multiple projections contribute to olfaction, so multiple challenges may arise in the generation of responses to an odor identification task.

While we only studied a 12-item version of this test, our results are consistent with the neuroscience of odor identification. Our Rasch model results suggested just three factors explaining at least 5% of the variability in this test; thus future modeling of more complex networks supporting odor identification, even with a greater number of items, should not be much more complex than three factors. Although we did find evidence of a single “real” factor, the test’s unidimensionality (30%) is so low that it is likely to “…threaten the interpretation of item and person estimates” (Smith, 2002, p. 206) that are derived from this test.

Two types of olfactory ability are frequently tested in a clinical setting (see Saive et al. 2014 for extensive review of the varieties of experimental paradigms to assess olfaction and associated neurological systems). One is the *detection* of a smell, which is assessed using a psychophysical tests designed to estimate the threshold amount of an odor required for the subject to detect that it is present – typically a single odor. This is a complex test to administer, cannot be given to more than one person at a time, and gives a very specific type of information about detection thresholds. The other type of test is of the *identification* (recognition and/or naming) of odors. In this task, participants or patients are asked to select the name for the smell with which they are presented from a four-choice forced alternative design (Hummel et al. 2011; see also Doty et al. 1995), or to determine whether a stimulant odor is more or less similar to a target odor (see Li et al. 2010). An identification or recognition task is simpler to administer (and score) than detection tasks, and recent versions of the four-choice alternative odor identification task have been presented on paper, making these much easier to administer within large cohorts – including the epidemiologic studies from which the data we analyzed for this paper were drawn. However, our analyses strongly suggest that the odor identification test does not give useful, or reliable, or invariant, information about olfaction (or cognition), so if there is an alternative type (not version) of test, it should be considered.

These negative results for the odor identification test are important and replicated findings that this test, although it has very high face validity, cannot be considered a reliable measure for the integrity of olfaction, olfactory cortex, general neuropathology, or even odor identification. It certainly cannot be used as a proxy for cognitive function and particularly not for change over time. This work also serves to underscore the fact that one cannnot depend on face validity alone – even if it is “supported” by classical estimates or characterizations of test-retest reliability (e.g., Doty et al. 1995; see also Yeshurun & Sobel, 2010). Our evidence suggests this test does *not* measure what it is perceived or believed to measure. Although it has very high face validity, the smell identification test is not a reliable test of olfaction, the integrity of olfactory cortex, general neuropathology, or even odor identification – *whatever it is* that the single ‘core’ construct representing the 30% of variability that was consistently found represents, it is not being measured well. The study is limited in that we did not include a formal, valid, and reliable assessment that actually measures olfactory cortex or odor identification. However, this limitation does not undermine the observation that multiple methods, applied to a very large sample, all converged on the same results.

In the best light, the 12-item odor identification test is extremely prone to error (roughly 70% noise), is not invariant across time or persons, and has a 30% signal that is actually global cognitive functioning (*not* olfaction). It can therefore not be utilized or conceptualized as a proxy for olfactory cortical (or other neuroarchitectural) integrity. It certainly should not be used as a proxy for cognitive function, nor should it be used for change over time.

## Data Availability

https://www.radc.rush.edu/docs/studyPublications.htm?studyName=MAP

## Declarations

### Ethics approval and consent to participate

all participants gave informed consent to participate in the longitudinal cohort study from which the data were derived. This consent included future research using their anonymized data.

### Consent for publication

NA

### Availability of data and material

The data that support the findings of this study are available from the Rush Alzheimer’s Disease Center, Chicago, IL (https://www.radc.rush.edu/docs/studyPublications.htm?studyName=MAP). Restrictions may apply to the availability of these data, which are available from the Center upon reasonable request and with permission of the Rush Alzheimer’s Disease Center.

### Authors’ contributions

RET and FY contributed equally to all parts of the manuscript, including all analysis and writing the manuscript. Both authors read and approved the final manuscript.

## Acknowledgements

The authors thank the participants of the Rush Memory and Aging Project for their invaluable contributions. We thank David Bennett, MD (RADC PI); Charlene Gamboa, MPH; Tracy Colvin, MPH; Tracey Nowakowski, Barbara Eubeler, and Karen Lowe-Graham, MS, for study recruitment and coordination, and John Gibbons, MS and Greg Klein for data management, and the staff of the Rush Alzheimer’s Disease Center.

The authors thank the participants of the Rush Memory and Aging Project for their invaluable contributions. We thank David Bennett, MD, Charlene Gamboa, MPH; Tracy Colvin, MPH; Tracey Nowakowski, Barbara Eubeler, and Karen Lowe-Graham, MS, for study recruitment and coordination, and John Gibbons, MS and Greg Klein for data management, and the staff of the Rush Alzheimer’s Disease Center. This research was supported in part by National Institute on Aging Grants [R01AG22018, R01AG17917, P30G10161], and the Illinois Department of Public Health.

